# Hypertension Beyond the Clinic: Patient Perspectives on Self-Management in a Resource-Limited Urban Setting

**DOI:** 10.1101/2025.04.28.25326544

**Authors:** Pius M Temba, Ruchius Philbert, Ashabilan A Ebrahim, Hassan Juma Rusobya, Fredirick L Mashili

## Abstract

**Background:** Uncontrolled hypertension remains a major contributor to morbidity and mortality in low- and middle-income countries (LMICs), particularly in Sub-Saharan Africa. While progress has been made in the diagnosis and detection of hypertension in Tanzania, effective long-term management continues to be hindered by challenges in patient self-management. Given that hypertension control largely depends on patients’ daily practices and adherence to lifestyle and treatment recommendations, understanding their experiences is essential. This study aimed to explore the lived experiences and perspectives of hypertensive patients regarding self-management practices in selected referral hospitals in Dar es Salaam, Tanzania.

**Methods:** This hospital-based qualitative study explored patients’ strategies and challenges related to hypertension self-management in selected regional referral hospitals in Dar es Salaam, Tanzania. Eleven participants with a diagnosis of hypertension for at least two years were purposively recruited from three facilities—Mwananyamala (n=5), Amana (n=3), and Temeke (n=3). Data were collected through in-depth, face-to-face interviews conducted in Swahili using a semi-structured guide. Interviews were audio-recorded, transcribed verbatim, and translated into English. Thematic analysis was conducted using an iterative process to identify emerging patterns and key themes. Ethical approval was obtained from the relevant institutional review board, and all participants provided written informed consent.

**Results:** Thematic analysis identified three main themes related to hypertension self-management: (1) Barriers to self-management, including limited knowledge, financial constraints, and lack of supportive systems; (2) Strategies to support self-management, such as family support, accessing nearby facilities for blood pressure monitoring, homegrown solutions for healthy eating, utilizing available spaces for physical activity, and seeking health information from providers or informal sources; and (3) Experiences with healthcare access, involving patient-provider relationships, transportation challenges, and prolonged waiting times. These findings highlight the complex interplay between individual agency, family dynamics, healthcare systems, and socioeconomic factors in shaping hypertension self-management practices.

**Conclusion:** This study highlights the importance of context-specific, patient-informed approaches to strengthen hypertension self-management in resource-constrained urban settings. The insights gained point to the need for integrated interventions that go beyond clinical care— addressing educational, social, and structural factors that shape patients’ daily management efforts. Empowering patients through tailored support systems and improving health service delivery are critical steps toward more effective and sustainable hypertension control.

## INTRODUCTION

There is accumulating evidence indicating an increasing burden of chronic non communicable diseases in the Sub Saharan African regions (12). Hypertension which is both a major risk factor and a common driver for cardiovascular and cerebrovascular diseases, account for 52.5% of all strokes in Africa (4). Globally, hypertension leads to 7.5 million cases which shares about 12.8% deaths recorded (7). Hypertension is often referred to as the “ silent killer” because most people are unaware; they have the condition in its early stages. It is usually detected when blood pressure is measured or after a serious medical complication occurs, such as a heart attack, stroke, or chronic kidney disease (20).

Although the detection and control of hypertension are improving in many countries, ad herence to medication and lifestyle changes remains low, making uncontrolled hypertension common (18). With the parallel rise in other NCDs an added burden to the already overstretched health care systems in SSA places significant strain on national budgets. Furthermore, the economic impact of NCD’s due to decreased productivity as a result of disability and death cannot be understated (13). In light of these challenges, it is clear that there is a need for management support to alleviate the burden of NCD’s. One form of intervention that has been used in recent years is self-management. This involves empowering individuals living with NCDs to take treatment in their own hands and enables patients to better manage their condition (19). Effective self-management is influenced by several factors such as the patients’ socio-economic status, their health literacy and access to care.

As self-management is primarily a patient-driven endeavor, their views must be taken into account. Despite this, data on patients’ perspectives on self-management in terms of challenges and strategies employed by patients is limited, especially in developing countries such as Tanzania. Sub Saharan Africa, with its social economic challenges present a unique entity that can potentially impact self management practices. These are often missed in quantitative studies given that most quantitative tools currently used were developed using studies mostly conducted in high income countries (18). Qualitative inquiry is particularly suited to uncover the lived experiences, contextual barriers, and complex coping mechanisms that structured surveys may overlook. These insights are critical not only for informing patient-centered interventions but also for guiding the development of future quantitative studies that can measure and validate the themes identified. Therefore, the aim of this study was to explore the challenges/barriers, experiences and strategies employed by patients towards hypertension self-management practices.

## MATERIALS AND METHODS

### Study design and setting

This was a hospital based qualitative study aimed at exploring the barrier and strategies related to hypertension self-management practices. The study was conducted at three regional referral hospitals in Dar Es Salaam, Tanzania: Temeke, Amana, and Mwananyamala. These hospitals host outpatient hypertension clinics, serving patients referred from lower-level facilities within their administrative areas. Clinics are staffed by at least one medical doctor and several nurses, providing diagnostic, follow-up and health promotion services including education, counseling, medication refills, and referrals when necessary.

### Study participant recruitment

Participants were purposively selected based on their experience with hypertension management. Inclusion criteria were: having a confirmed hypertension diagnosis for at least 12 months and attending the clinic for a minimum of six months. Recruitment was facilitated by unit in-charges, who introduced the research team and assisted in identifying eligible participants. The researcher then explained the purpose of the study and obtained informed written consent. Eleven participants were recruited for in-depth interviews, with data saturation reached after eight interviews; three additional interviews were conducted to confirm thematic redundancy.

### Data Collection

Face-to-face in-depth interviews were conducted between May and July 2020 in a private room near the outpatient clinic to ensure confidentiality. Interviews were conducted in Swahili by trained researchers with prior experience in qualitative research and a medical background in hypertension care. Each interview lasted between 30 and 60 minutes and was audio-recorded with participants’ consent. All interviews followed a semi-structured guide developed to explore barriers and self-management Strategies.

### Data processing and analysis

Audio recordings were transcribed verbatim in Swahili, and then translated into English for analysis. Data were analyzed thematically through an iterative process. After initial review of the first transcript, the research team met to discuss emerging concepts and refine the interview process. Line-by-line coding was first applied to one transcript collaboratively to develop an initial codebook. Subsequent transcripts were coded independently by team members using this codebook. Discrepancies in coding were resolved through team discussions, and themes were finalized by consensus. Microsoft Excel was used to organize and sort coded data into subthe mes and themes.

### Study Rigor

To enhance the trustworthiness of the study, we employed several strategies aligned with qualitative research quality criteria. Credibility was supported through prolonged engagement during interviews, daily debriefings among the research team, and iterative refinement of the interview guide. Dependability was ensured through consistent documentation of coding decisions and peer review of coded data. Confirmability was enhanced by maintaining an audit trail of analytical decisions and using team consensus to finalize themes. While member checking was not conducted due to logistical constraints, efforts were made to reflect participants’ voices accurately through verbatim transcription and contextual interpretation of responses.

### Ethical consideration and informed consent

This study was approved by the Muhimbili University of Health and Allied Sciences (MUHAS) Institutional Review Board (Ref. No. DA.282/298/01.C/). Permission was also obtained from the medical officers in charge of the participating hospitals. All participants provided written informed consent and were informed of their right to withdraw at any time. Anonymity was maintained through the use of unique identifier codes during data collection and analysis.

## RESULTS

### Characteristics of participants

The study purposefully recruited eleven participants attending hypertension follow-up clinics at three regional referral hospitals in Dar es Salaam. Among the participants, seven were males, and four were aged over 55 years. Most were married (n = 9), with the remainder widowed. Participants were distributed across three hospitals: five from Mwananyamala, three from Amana, and three from Temeke Regional Referral Hospital. All participants had been diagnosed with hypertension for at least two years.

### Thematic analysis

Thematic analysis identified three central themes reflecting barriers and experiences in hypertension self-management: (1) Barriers to self-management practices, (2) Strategies employed to support self-management, and (3) Experiences with healthcare service access. Each theme includes subthemes derived from participant narratives and supported with illustrative quotes.

Theme **1: Barriers to self-management practices**

#### Limited Knowledge on Self-Management

When asked about their understanding of hypertension self-management, participants revealed varying levels of awareness. While some had never encountered the term, others associated it with taking control of their health through lifestyle changes. Dietary management strategies mentioned included reducing salt, carbohydrates, and fatty foods, while increasing vegetables and fruits. Participants also described physical activity practices such as walking and stretching exercises, as well as weight management. One participant expressed:

> *“ It is the self-awareness and acceptance of taking full control of your disease, acknowledging the illness, and being ready to take initiatives such as engaging in exercise, reducing salt intake, and losing weight for better health (IDI Participant 05)*.*”*

#### Financial Barriers to Self-Management

While many participants reported medication adherence, financial constraints were a recurrent barrier. Even insured individuals noted that some medications were not covered, requiring out-of-pocket payments that were not always feasible. For example:

> *“ It’s not that I forget to take the medication*… *for example, they prescribed me with a month’s dose, and then I find that I don’t have the cash to pay for it. (IDI Participant 08)”*
>
> *“ Most of the prescribed medications require money to obtain, so it depends on whether I have the cash at that time. If I don’t have money, I have to wait until I can afford to purchase the medication. (IDI Participant 12)”*
>
> Additionally, limited financial means affected patients’ ability to maintain recommended diets and conduct routine blood pressure checks. As one participant explained:
>
> *“ If you want to consistently maintain a healthy meal plan, you need money. These diseases are for the rich, as they can afford to choose what they eat. (IDI Participant 01)*.*”*

#### Lack of Supportive systems for hypertension self-management

Participants highlighted a significant gap in the availability of supportive systems necessary for effective hypertension self-management. They expressed concerns over the insufficient health education provided by healthcare providers, noting that clinic appointments were primarily focused on medication refills rather than offering comprehensive support for managing their condition. Additionally, they emphasized on the absence of infrastructure to facilitate physical activities, as well as the lack of patient support groups where individuals could share experiences and motivate one another in their self-management journey. One participant stated;

> *“ We need a specific place for the hypertensive patients to exercise, with special trainer having understanding about type of exercise special for a person with hypertension (IDI Participant 08)*.*”*

Similarly, another respondent stated;

> *“ We are encouraged to exercise, but we have not been educated on the types of exercises we should be doing. We need a trainer to guide us. (IDI Participant 04)*.*”*

**Theme 2: Strategies Supporting Self-Management**

#### Family support in self-management

Participants described how family members played a pivotal role in supporting their self-management practices. This included financial assistance, reminders for medication, and shared participation in physical activity. For example:

> *“ For sure, there are no challenges. After I meet with the doctor and receive my prescribed medication, I call my brother, who sends me money to buy the pills. (IDI Participant 10)” “ Thanks to my husband, sometimes when he comes home from work, he takes me for a walk. (IDI Participant 08)”*
>
> *“ I have never forgotten to take my hypertensive medication because my husband usually reminds me. Since the medicines are expensive, I purchase them on a ten-day basis, and when they run out, I remind my husband to refill them. (IDI Participant 06)”*

#### Accessing Nearby Facilities for Blood Pressure Monitoring

Due to the high cost of personal monitoring devices and clinic visits, participants reported using nearby pharmacies or dispensaries for routine blood pressure checks. These were seen as affordable and convenient alternatives:

> *“ Mostly, I do my checkups at home, checking my blood pressure every week. There is a pharmacy near where I live, so I just pay 1*,*000 TSH for the checkup. (IDI Participant 07)”*

#### Homegrown Solutions for Nutrition and Cost-Saving

Several participants adopted innovative practices to improve their diet without incurring high costs. These included growing vegetables at home and raising poultry, which helped reduce reliance on market purchases.

> *“ I have a garden at home where I grow a variety of vegetables. It helps me reduce the cost of buying vegetables. (IDI Participant 05)”*

#### Utilizing available space for physical activity

Despite the lack of formal facilities, participants described engaging in physical activity using available community spaces or public roads. Activities included walking, stretching, and occasional gym visits.

> *“ I also usually go to the gym, where we do rope skipping, and stretching exercises; when you do it regularly your blood pressure tends to normalize. (IDI Participant 11)”*
>
> *“ I regularly do exercise every evening, walking, running then walking again when I feel tired. (IDI Participant 03)”*

#### Access to health information

Participants acknowledged receiving health education during clinic visits and expressed appreciation for advice from healthcare providers. Others reported seeking additional information through online platforms or family members with similar health conditions.

> *“ Every Wednesday is scheduled for health education, so we usually come to the clinic for classes whereby doctors and nurses teach us about different health topics related to our illness. (IDI Participant 03)”*
>
> *“ My father is also hypertensive, and he’s the one who advises me on the lifestyle changes I need to adopt. (IDI Participant 06)”*

**Theme 3: Experiences with health care access**

#### Patient-provider relationships

While many participants described positive relationships with healthcare providers, characterized by good communication and trust, others expressed frustration over short consultations and limited time to discuss concerns.

> *“ We communicate well with caregivers; they are always ready to help you whenever you have a problem. (IDI Participant 02)”*
>
> *“ I ask him [the doctor], and then he will respond to my questions regarding my illness, but sometimes you’re told, ‘Please go, other patients are waiting*.*’ So, I feel like I don’t have enough time to express all my concerns. (IDI Participant 09)”*

#### Transportation to health facility

Transportation costs and distance were frequently cited challenges. Some participants walked to clinics, when possible, while others spent considerable time and money commuting.

> *“ I just walk to the clinic; for me, I consider it part of my exercise. I only take a daladala when the day is very sunny, and it may cost me up to 1*,*000 TSH. It can take me almost an hour to get here. (IDI Participant 12)”*
>
> *“ I do take four buses to reach the hospital as I reside in Lugoba, Chalinze, so I almost use more than four hours to reach here, I almost spend around 10*,*000 TSH as transport fare. (IDI Participant 05)”*

#### Service waiting time

Long waiting times at clinics were commonly reported. Participants expressed concerns over the time spent waiting relative to the short consultation period.

> *“ Sometimes I arrive here at 7 a*.*m*., *but I end up waiting until 11 a*.*m. to see the doctor. When I see him, it’s usually for a very short time, often less than ten minutes, which is too short for me to explain everything. (IDI Participant 07)”*

## DISCUSSION

This study aimed to explore the challenges and strategies related to hypertension self-management among patients attending regional referral hospitals in Dar es Salaam. The findings revealed that limited understanding of hypertension self-management, lack of supportive systems, and financial constraints significantly hindered self-management practices. Patients adopted several strategies to cope, including utilizing nearby health facilities, engaging in available infrastructure for physical activity, relying on homegrown solutions for affordable, nutritious food, and drawing on family support to manage their condition.

Access to medication and healthcare services emerged as a major barrier to effective hypertension self-management. Participants in this study experienced substantial financial constraints, particularly when medications were not covered by insurance. These financial barriers often delayed medication acquisition, exacerbating the challenges of managing their condition. This reflects broader systemic issues in healthcare accessibility, including gaps in insurance coverage and high out-of-pocket expenses, consistent with other studies in similar low-resource settings (21, 8).

The lack of supportive systems for self-management was another critical challenge. Participants highlighted the absence of community infrastructure such as exercise spaces and trained professionals knowledgeable about hypertension-specific physical activity. These findings echo previous research underscoring the need for structured community-based resources to support chronic disease self-management. (3, 6).

Family support was a key facilitator of adherence and engagement in self-management practices. This included financial help, reminders to take medications, and encouragement to engage in physical activity. The role of caregivers and social networks in supporting chronic disease management is well-established (17), and these findings reinforce the need to incorporate family-based strategies into hypertension care models, especially in resource-limited environments. Participants also reported using nearby pharmacies and dispensaries for affordable blood pressure monitoring, minimizing transportation costs. This aligns with studies highlighting the role of decentralized, community-based services in improving chronic disease outcomes in underserved populations. (15).

Self-sufficiency strategies such as growing vegetables and raising poultry were adopted to improve dietary quality without incurring high food costs. These practices align with public health recommendations promoting local, sustainable food sources to reduce non-communicable disease risk in low-income populations. (5).

The patient-provider relationship also played a central role in shaping participants’ experiences. While many praised the communication and professionalism of healthcare workers, some participants expressed dissatisfaction with brief consultations focused primarily on medicatio n refills. This highlights a need for more holistic, patient-centered care that includes education, counseling, and space for patient concerns. (14).

Transportation challenges and clinic accessibility were additional barriers. Participants noted the burden of long distances, high transport costs, and extended waiting times at clinics, all of which influenced their ability to adhere to medical advice. These barriers are consistent with findings from other African settings and further emphasize the need for decentralized, community-based interventions. (9, 17).

While this study offers valuable insights into the lived experiences of patients managing hypertension in an urban African context, it is not without limitations. The qualitative design, while well-suited for capturing depth and context, limits the generalizability of findings. Additionally, responses may have been shaped by social desirability or participants’ comfort in discussing sensitive issues. Nevertheless, the rigorous thematic analysis and participant-centered approach offer a strong foundation for future research. These findings may inform the development of targeted, domain-specific quantitative tools and larger-scale interventions that address both structural and behavioral dimensions of chronic disease self-management.

Overall, this study underscores the multifaceted barriers to hypertension self-management in Dar es Salaam, highlighting the need for improved patient education, financial support, and enhanced community and healthcare infrastructure. Addressing these challenges through policy, family-centered approaches, and systems-level reforms will be essential to improving hypertension control and reducing cardiovascular risk in Tanzania and similar settings.

## CONCLUSION

This study underlines the multifaceted challenges individuals with hypertension face in managing their condition, from financial barriers to the accessibility of healthcare services and self-management resources. While many participants actively engaged in self-management practices, they often encountered obstacles related to the affordability of medications, availability of appropriate facilities, and the support provided by healthcare systems. Addressing these challenges through improved access to affordable medications, better infrastructure for physical activity, and more comprehensive healthcare services could significantly enhance the effectiveness of hypertension self-management. Moreover, leveraging family support and community resources remains crucial in helping individuals manage their condition effectively, particularly in low-resource settings.

## Data Availability

All data produced in the present work are contained in the manuscript

## Author contributions

All authors contributed to conceptualization of the study, data collection and manuscript development equally.

## Data sharing statement

We will provide any primary data access when requested

